# Leaving against medical advice as a signal of unmet care needs in adult sickle cell disease hospitalizations

**DOI:** 10.64898/2026.03.20.26348715

**Authors:** Anna Zhilkova, Kenneth Rivlin, Jenai Jackson, Jeffrey Glassberg, Brittany McCrary, Antonia Eyssallenne

## Abstract

**Importance:** Sickle cell disease (SCD) affects approximately 100,000 people in the United States, causes life-threatening complications, and shortens life expectancy by decades. Adults with SCD routinely encounter undertreated pain, provider bias, and structural barriers in hospital settings.

**Objective:** To describe patterns of leave against medical advice (LAMA) among adults hospitalized for SCD.

**Design, Setting, and Participants:** Retrospective analysis of inpatient discharge records among adults ages 18 and older in New York City hospitals, 2022-2023, hospitalized for SCD or any reason.

**Main Outcomes and Measures:** The primary outcome was hospital-level LAMA, measured by crude rates and rates adjusting for patient characteristics using Bayesian hierarchical models. The secondary outcome was 30-day all-cause readmissions, stratified by LAMA status.

**Results:** LAMA discharges comprised 14% of SCD hospitalizations and 4% of all-cause hospitalizations. Adjusted hospital-level SCD LAMA ranged from under 5% to 30% (IQR: 10–20%) and was higher than all-cause LAMA in most facilities. Crude SCD LAMA rates exceeded 30% in several hospitals, including those with more than 100 SCD hospitalizations during the study period. Patients with 10 or more SCD hospitalizations accounted for 40% of total SCD volume. Sensitivity analyses accounting for this concentration showed attenuated but persistent variation in SCD LAMA. Over 50% of SCD LAMA discharges were followed by a 30-day readmission compared to 38% of non-LAMA discharges. LAMA was associated with higher adjusted odds of readmissions in both SCD and all-cause hospitalizations.

**Conclusions:** Our findings challenge the assumption that patients are solely responsible for early departures. Leaving against medical advice should be monitored as a signal of unmet care needs in SCD.

## Introduction

Sickle cell disease (SCD) is the most common inherited blood disorder in the United States (US), affecting about 100,000 people (Hassell 2010). Most people in the US with SCD are of African ancestry, although the condition can also be found among individuals with Mediterranean, Middle Eastern, and South Asian ancestry reflecting the geographic distribution of the sickle cell mutation along historic malaria belts.

People with SCD experience life-threatening complications that require hospital care and can shorten life expectancy by decades (Lubeck 2019). These complications include painful vaso-occlusive episodes, acute chest syndrome, stroke, and progressive organ failure. In hospital settings, people with SCD routinely encounter providers who question the severity of their pain, label them as drug-seeking, and delay opioid administration as compared to patients with other painful conditions such as long-bone fractures or renal colic (Haywood 2010, Haywood 2013, Lazio 2010). Such experiences may stem from systemic- and provider-level factors beyond patient control, including limited SCD training, outdated clinical protocols, implicit bias, stigma around pain management, and structural racism in healthcare (Bulgin 2018, Glassberg 2013). As observed by Power-Hays and McGann (2020), “there may be no population of patients whose healthcare and outcomes are more affected by racism than those with sickle cell disease.”

Hospitalizations for SCD deserve special attention because of exceptionally high rates of readmissions and other adverse outcomes. Over 30% of adult SCD hospitalizations result in a 30-day readmission – the highest of any diagnosis and nearly triple the national average among US adults (Goel 2025, Jiang 2024). SCD hospitalizations are also about four times more likely to result in patients leaving against medical advice (LAMA) (Fingar 2019). The reasons for LAMA may be wide-ranging. But for patients presenting with SCD complications, these departures may reflect well-documented frustration with undertreated pain (Haywood 2010) and broader issues with health systems that leave patient needs unmet and clinicians insufficiently supported to deliver effective care. Together, these patient reports and outcomes suggest unaddressed quality gaps in acute care for SCD.

Common hospital performance measures often overlook or exclude LAMA discharges (YNHHSC/CORE 2024). As a result, hospitals receive no explicit feedback on this outcome. It also implies that patients who leave are a homogeneous group largely responsible for their own departures. Emerging perspectives, however, challenge this assumption and argue that LAMA reflects gaps in health care delivery (Ambasta 2020, Holmes 2024, Rivlin 2025).

In this study, we describe patterns of LAMA using population-level hospital discharge records from New York City – home to one of the largest populations of people living with SCD in the US. Building on prior work describing hospital-level variation in unadjusted SCD LAMA rates (Rivlin 2025), we examine variation after adjusting for patient characteristics and assess whether LAMA discharges are associated with higher risk of 30-day readmissions.

## Methods

### Data source

We used inpatient hospital discharge data from SPARCS (Statewide Planning and Research Cooperative System), an all-payer system maintained by the New York State Department of Health. Each record represents a hospital discharge and includes patient demographics, admission characteristics, diagnoses, procedures, and facility information. Some facilities, such as federal and psychiatric hospitals are not captured by the system.

### Study setting and population

We extracted records for adults 18 years and older discharged from New York City hospitals from January 1, 2022 through December 31, 2023. For descriptive LAMA analyses, index hospitalizations were defined as discharges occurring January 1, 2022 through November 30, 2023. December 2023 discharges were retained only to flag readmissions. Hospitalizations for SCD were identified using ICD-10-CM codes D57.x in the principal diagnosis, excluding D57.3 (sickle cell trait). We excluded records with missing information about admission date, discharge date, or patient IDs required to track readmissions. Hospitals with fewer than 20 SCD hospitalizations were excluded to ensure more stable estimates. A flow diagram is provided in eFigure 1.

### Primary outcome

The primary outcome was hospital-level LAMA rates. LAMA events were identified using the patient disposition code 07 (“Left against medical advice”). Adjusted LAMA rates were calculated as the ratio of each hospital’s predicted LAMA cases to its expected LAMA cases multiplied by 100 (YNHHSC/CORE 2024, O’Brien 2016).

Predicted and expected LAMA cases were obtained using Bayesian hierarchical logistic regression models with a hospital-level random intercept to stabilize estimates for hospitals with few events. We fit models to predict LAMA (yes/no) with the following binary patient-level clinical predictors: sex (female), age group (18–29, 30–49, 50–64, ≥65), emergency department (ED) admission, severity of illness present on admission (1-4) based on APR-DRG (All Patient Refined Diagnosis Related Groups), and Elixhauser comorbidities without HIV. Further modeling details are provided in the Supplement.

We performed two sensitivity analyses for hospital-level SCD LAMA rates: by incorporating a crossed patient-level random intercept to account for correlation among patients with multiple SCD hospitalizations and by excluding patients with 10 or more hospitalizations.

### Secondary outcome

The secondary outcome was all-cause 30-day hospital readmission, stratified by LAMA status. We used Agency for Healthcare Research and Quality (AHRQ) methodology to identify hospital readmissions with several modifications, including exclusion of planned hospitalizations and a modified approach to identifying transfers. Additional details are provided in the Supplement.

We also examined the association between leaving against medical advice (LAMA) at the index discharge and 30-day all-cause readmission. Readmission (yes/no) was modeled at the discharge level using hierarchical logistic regression models incorporating a hospital-level random intercept. The main parameter of interest was the adjusted odds ratio (aOR) for LAMA. Model 1 adjusted for patient characteristics described above. Model 2 included all model 1 predictors and added the following demographic predictors: race/ethnicity (Hispanic, Asian or Pacific Islander, Black, White, or Other), insurance type (Medicaid, Medicare, commercial, self-pay), and New York City neighborhood (Community District) fixed effects. Under 1% of records had missing data on predictors and were excluded.

Analyses were conducted using R version 4.3 and Stata 16.

### Ethics

This study was approved by the New York City Department of Health and Mental Hygiene’s Institutional Review Board as exempt human subjects research and complied with the Strengthening the Reporting of Observational Studies in Epidemiology (STROBE) guidelines.

### AI use disclosure

We used Claude Opus 4.6 (Anthropic) and ChatGPT 5 (OpenAI) to review code, methods, and manuscript. No patient data was uploaded. We take full responsibility for the content and any errors.

## Results

There were 7,939 SCD index hospitalizations and 1,358,810 all-cause index hospitalizations among adults in New York City hospitals from January 1, 2022 through November 30, 2023 (Table 1). LAMA discharges comprised 14% of SCD hospitalizations and 4% of all-cause hospitalizations. Adjusted hospital-level SCD LAMA rates ranged from under 5% to 30% (IQR 10-20%) (Figure 1). Crude rates exceeded 30% in several hospitals, including hospitals with more than 100 SCD hospitalizations during the study period (eTable 1). Adjusted SCD LAMA rates exceeded all-cause LAMA rates in most facilities.

**Table 1.**
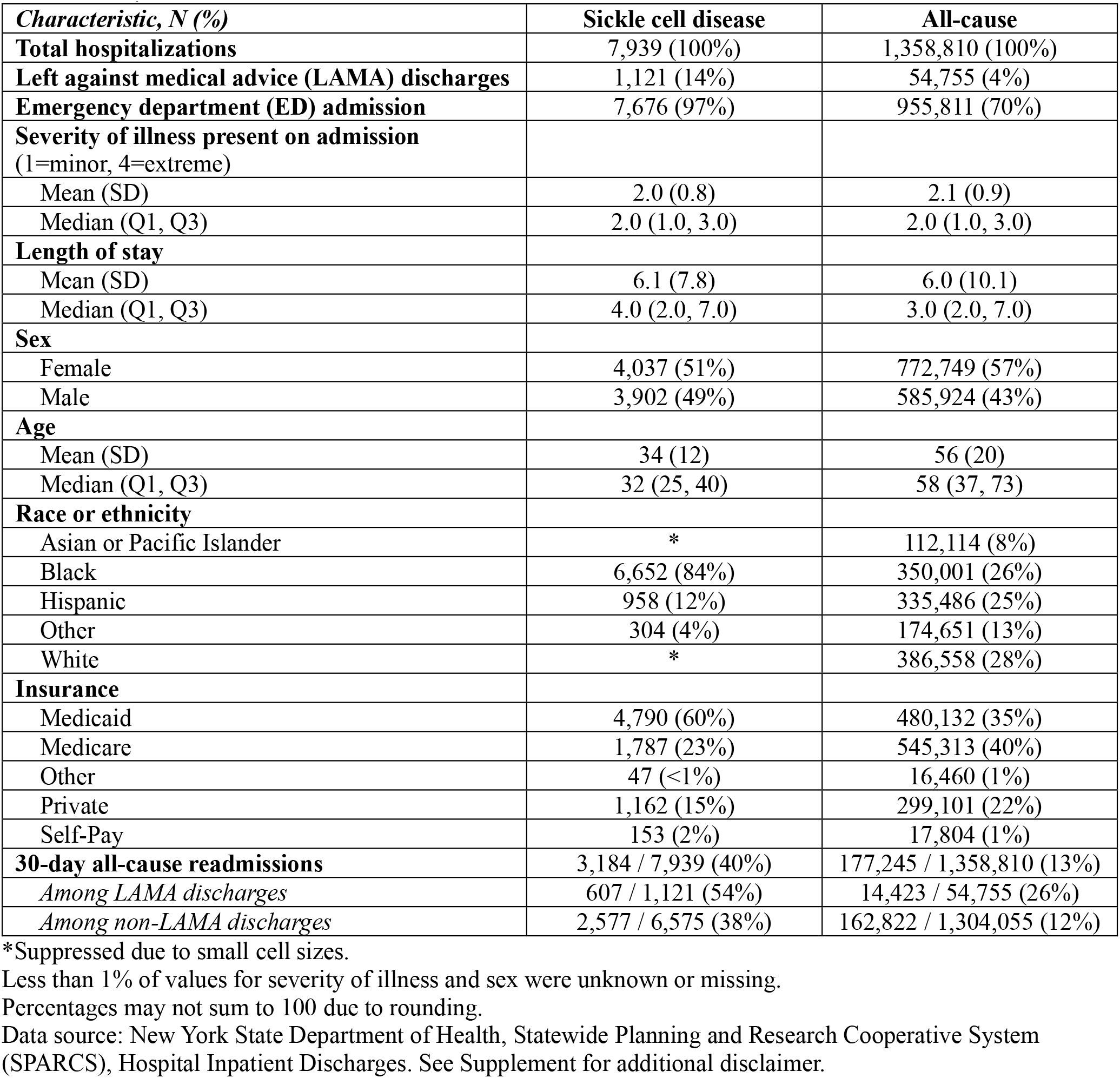
Sickle cell disease and all-cause index hospitalizations among adults ages 18 and older, January 1, 2022-November 30, 2023.

**Figure 1.**
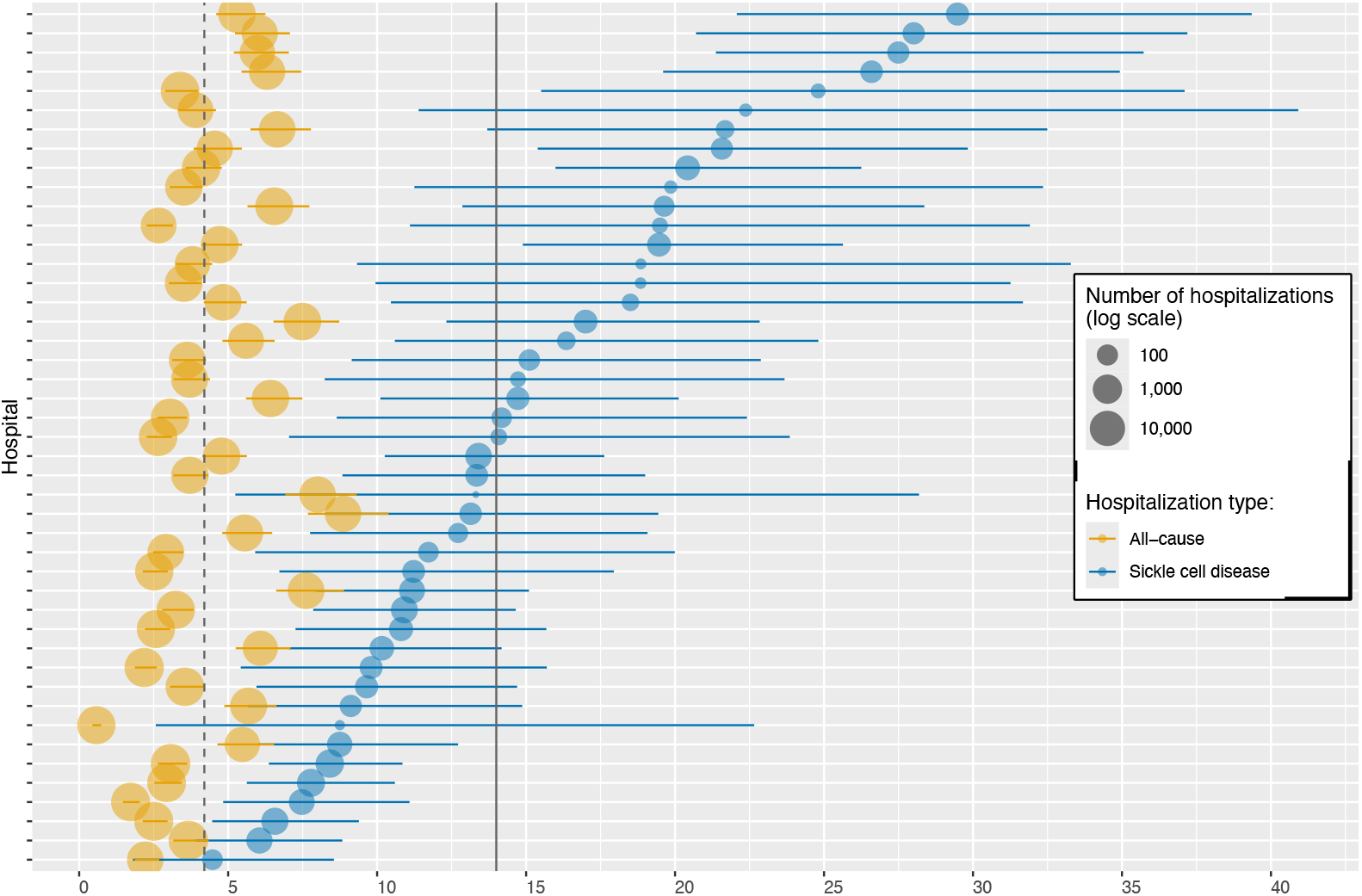
Hospital-level leave against medical advice (%) for sickle cell disease and all-cause adult hospitalizations, January 1, 2022-November 30, 2023 *Facilities (N=45) sorted by adjusted SCD LAMA* Solid line = SCD hospitalizations average; dashed line = all-cause hospitalizations average. LAMA adjusted for patient characteristics: sex (female), age group (18–29, 30–49, 50–64, ≥65), emergency department (ED) admission, severity of illness present on admission (1-4) based on APR-DRG (All Patient Refined Diagnosis Related Groups), Elixhauser comorbidities excluding HIV, and a hospital random intercept. Error bars indicate 95% credible intervals. Hospitals with fewer than 20 adult SCD hospitalizations were excluded from the analysis. Data source: New York State Department of Health, Statewide Planning and Research Cooperative System (SPARCS), Hospital Inpatient Discharges. See Supplement for additional disclaimer.

Multi-visit patients contributed to a substantial share of all SCD hospitalizations. Among over 2,000 unique patients with an SCD hospitalization, about 7% had 10 or more SCD hospitalizations. These patients accounted for 40% of the total SCD volume during the study period. By comparison, under 1% of patients in the all-cause group had 10 or more hospitalizations and contributed to about 3% of the total volume (Table 2).

**Table 2.** Distribution of unique patients contributing to index sickle cell disease and all-cause hospitalizations, January 1, 2022-November 30, 2023.

|  | **Sickle cell disease** | **All-cause** |
| --- | --- | --- |
| **Unique patients with:** |  |  |
| 1-3 hospitalizations | 1,766 (76%) | 882,114 (95%) |
| 4-9 hospitalizations | 381 (16%) | 40,917 (4%) |
| 10+ hospitalizations | 166 (7%) | 3,173 (<1%) |
| **Total unique patients** | **2,313 (100%)** | **926,204 (100%)** |
| **Hospitalization volume among unique patients with:** |  |  |
| 1-3 hospitalizations | 2,602 (33%) | 1,106,925 (81%) |
| 4-9 hospitalizations | 2,161 (27%) | 207,581 (15%) |
| 10+ hospitalizations | 3,176 (40%) | 44,304 (3%) |
| **Total hospitalizations** | **7,939 (100%)** | **1,358,810 (100%)** |

Percentages may not sum to 100 due to rounding.
Data source: New York State Department of Health, Statewide Planning and Research Cooperative System (SPARCS), Hospital Inpatient Discharges. See Supplement for additional disclaimer.

Adjusted SCD LAMA rates by hospital were modestly compressed using a sensitivity model that added a crossed patient-level random intercept to account for correlation among multi-visit patients (IQR 11-19% vs. 10-20% in the baseline model) (eTable 1). In the second sensitivity analysis, excluding patients with 10 or more SCD hospitalizations reduced adjusted hospital-level variation (IQR 8-14% vs. 10-20%) (eTable 1). The overall SCD LAMA rate was reduced from 14% to 11%, while all-cause LAMA remained at 4% after exclusion of patients with 10 or more hospitalizations.

The 30-day readmission rate was 40% for SCD hospitalizations and 13% for all-cause hospitalizations (Table 1). A 30-day readmission occurred in over 50% of SCD LAMA hospitalizations and 38% of non-LAMA SCD discharges. After adjusting for patient characteristics, LAMA was associated with a higher risk of a 30-day readmission among sickle cell disease hospitalizations (adjusted OR= 1.63, CI95 = 1.39-1.91) and among all-cause hospitalizations (adjusted OR = 2.10, CI95 = 1.96-2.26). Consistent results were obtained for models that added demographic and neighborhood predictors (eTables 3-4).

## Discussion

Adult SCD hospitalizations were more than three times more likely to result in patients leaving against medical advice, and over half of these SCD LAMA discharges were followed by a 30-day readmission. We found wide variation in SCD LAMA across hospitals that persisted after adjusting for patient characteristics under different models. These findings challenge the assumption that patients are solely responsible for early departures.

Two other findings are noteworthy. First, our sensitivity analysis revealed that SCD LAMA, but not all-cause LAMA, is substantially driven by a small number of patients with 10 or more hospitalizations. The needs of multi-visit patients with SCD are crucial to understand and address. Second, readmission rates were elevated even among non-LAMA SCD discharges and also warrant attention.

Hospitals can take meaningful steps to strengthen how they care for people with SCD. Readily implementable steps include root cause analysis about why patients leave, aligning protocols to reflect clinical guidelines, and routine monitoring of health care inputs and outcomes (Table 3). Investing in resources such as social workers can strengthen care coordination and support patients with complex needs, including those of multi-visit patients. Public health departments can support these efforts by reporting hospital outcomes and collaborating with health systems and community partners to strengthen care.

**Table 3.**
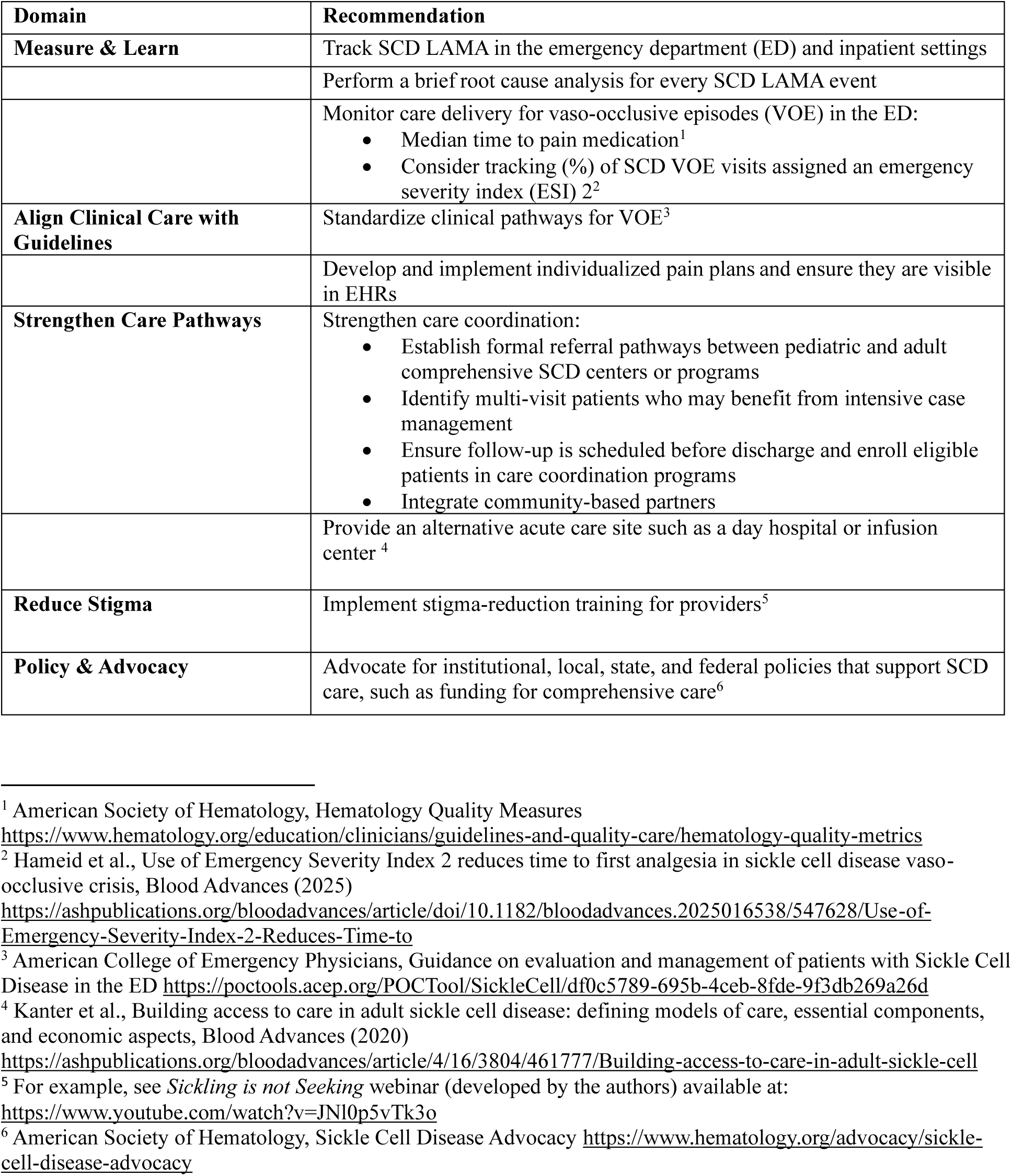
Recommendations to strengthen care for sickle cell disease in hospital settings.

### Limitations and future research directions

This analysis has several limitations. First, administrative data has limitations about quality of documentation. This affects our ability to capture important confounders such as disease severity. Additionally, if some LAMA events are reported as routine discharges, then our results would underestimate true LAMA rates. Second, our baseline model with patient characteristics and a hospital-level random intercept is similar to established approaches for standardizing hospital outcomes such as those used by the Centers of Medicare & Medicaid (CMS), but it does not account for correlation among patients with multiple hospitalizations. Our sensitivity analysis showed that addressing this through addition of a patient random intercept modestly attenuated SCD LAMA variation. Hence, the baseline model overstates the hospital-level SCD variation. On the other hand, the sensitivity model may understate variation attributable to the hospital if a patient’s likelihood of LAMA is itself shaped by past health care experience. Substantial hospital-level variation in SCD LAMA persisted using different models. Adjusted rates under different modeling assumptions and crude rates may be informative to consider together when assessing hospital outcomes. Third, adjusted hospital-level rates rely on indirect standardization - an established approach when incorporating multiple patient predictors, but which may not fully account for unmeasured differences in patient complexity across facilities (Shahian 2008). Fourth, this study looks at outcomes in an urban area with several academic medical centers, and our findings may not generalize to other healthcare markets.

Future research directions include replicating the analysis in different health care markets, characterizing emergency department departures, and exploring other conditions with high LAMA rates that may indicate gaps in health care delivery. Examining health care inputs such as timing of pain management – available in EHRs but not in administrative data – may reveal modifiable predictors of LAMA. Lastly, causal designs that isolate patient and hospital factors when assessing variation in outcomes such as readmissions can be extended to LAMA as well (Chandra 2023, Doyle 2019, Krumholz 2017).

## Data Availability

The underlying data used in this study cannot be made publicly available due to patient privacy.

