## Supplement for "Leaving against medical advice as a signal of unmet care needs in adult sickle cell disease hospitalizations"

**eFigure 1:** Inclusion and exclusion criteria for index hospitalizations, January 1, 2022-November 30, 2023[1]

**New York City inpatient hospital discharges, 2022-2023**

- *Includes NYC hospitals or NYC residents discharged from hospitals in NY state outside the city*

**2,085,674** all-cause hospitalizations**,** including **11,226** SCD hospitalizations

↓

**Ages 18 and older in NYC hospitals**

**1,607,929** all-cause hospitalizations, including **8,568** SCD hospitalizations

↓

**Complete records**

- *Excludes records with missing data on admission date, discharge date, patient ID*

**1,550,493** all-cause hospitalizations, including **8,387** SCD hospitalizations

↓

**Index hospital discharge January 1, 2022-November 30, 2023**

- *Excludes in-hospital deaths, discharges to a hospice, December 2023 discharges (due to insufficient follow-up time for readmissions)*

**1,425,699** all-cause hospitalizations, including **7,983** SCD hospitalizations

↓

**Hospitals with 20 or more SCD index hospital discharges**

**1,358,810** all-cause hospitalizations including **7,939** SCD hospitalizations

*Final study sample*

[1] December 2023 discharges were not eligible as index hospitalizations due to insufficient 30-day follow-up, but were retained in the data to ascertain readmissions occurring within 30 days of November 2023 index discharges.

### Modeling predicted and expected LAMA counts by hospital

We fit Bayesian hierarchical logistic regression models to obtain predicted and expected LAMA cases using the *rstanarm* R package version 2.32.1[1]. The following priors were assigned: N(0,1) for the coefficients of patient-level binary predictors, Student-t (3, 0, 5) for the intercept, and exponential(1) for the standard deviation of the hospital random intercept. Four Markov chain Monte Carlo (MCMC) chains were run for 3,000 iterations each, including 1,000 warm-up iterations, for a total of 8,000 post-warm-up draws from the posterior distribution.

For each draw from the posterior distribution, two sets of patient-level predicted probabilities of LAMA were computed: 1) by including the hospital's random intercept, representing the hospital's predicted performance, and 2) by setting the hospital random intercept to zero, representing the expected performance of an average hospital with the same case mix based on measured patient characteristics. Patient-level probabilities were summed for each hospital to obtain posterior distributions of predicted and expected LAMA counts. Hospital-level adjusted LAMA rates were reported as the posterior median (point estimate) and the 95% credible interval (2.5th and 97.5th percentiles of the posterior distribution). Analyses were performed for SCD and all-cause hospitalizations.

For the sensitivity analysis that incorporated a crossed patient-level random intercept into the baseline model for SCD hospitalizations, we tested two different priors using an exponential distribution (implemented below). Both produced similar results of hospital-level LAMA rates. We did not perform the sensitivity analysis for all hospitalizations due to computational constraints. However, our results suggested that the distribution of patients with multiple all-cause hospitalizations is less of a concern than for SCD.

We performed basic diagnostics of MCMC convergence that did not indicate major issues.[2]. R-hat values were at or below 1.01, bulk and tail effective sample size (ESS) exceeded 400 for all parameters, and no divergent transitions were found.

Sample code for fitting the models:

```
formula1 <- as.formula(paste("lama ~ female + agegroup + ed + soi_poa + ",
paste(elix_dummies, collapse = " + "), " + (1 | fac_id)"))

# Sensitivity model
# formula2 <- as.formula(paste("lama ~ female + agegroup + ed + soi_poa + ",
# paste(elix_dummies, collapse = " + "), " + (1 | fac_id) + (1 | id1) "))

fit_stan <- stan_glmer(
  formula1,
  data    = data1,
  family  = binomial(link = "logit"),
  prior   = normal(0, 1), # for coefficients of pt binary predictors
  prior_intercept  = student_t(3, 0, 5), # for intercept
  prior_covariance = decov(shape = 1, scale = 1), # for SD of hosp random intercept
# priors for SD of hosp- AND pt-level random intercepts (note the order)
# prior_covariance = decov(shape = 1, scale = c(1,1)),
# less informative, less shrinkage for pt-level (note the order)
# prior_covariance = decov(shape=1, scale = c(1, 2.5)),
```

---

[1] Goodrich B, Gabry J, Ali I & Brilleman S. (2025). rstanarm: Bayesian applied regression modeling via Stan. R package version 2.32.1. https://mc-stan.org/rstanarm.
For up to date information about package defaults: https://cran.r-project.org/web/packages/rstanarm/rstanarm.pdf

[2] Vehtari A, Gelman A, Simpson D, Carpenter B, Bürkner PC. Rank-Normalization, Folding, and Localization: An Improved $\hat{R}$ for Assessing Convergence of MCMC (with Discussion). *Bayesian Anal*. 2021;16(2):667-718. doi:10.1214/20-BA1221. https://projecteuclid.org/journalArticle/Download?urlId=10.1214%2F20-BA1221

```
  chains  = 4,
  iter    = 3000,
  warmup  = 1000,
  adapt_delta = 0.95,
  cores  = min(4, parallel::detectCores() ),
  init = "0"
)
```

**Readmissions methodology**

We identified all-cause 30-day readmissions using a modified approach to Agency for Healthcare Research and Quality (AHRQ) methodology.[3] An index hospitalization was defined as any hospitalization that could be followed for 30 days and did not result in transfer to another acute care hospital. We excluded in-hospital deaths, discharges to hospice and December 2023 discharges from index hospitalizations (due to insufficient follow-up time).

We defined a readmission as a subsequent hospital admission to the same or different hospital within 30 days following the index discharge. We modified the AHRQ methodology in several ways. First, we limited readmissions to unplanned hospitalizations, defined as emergency, urgent, or trauma admissions. Second, we identified transfers as admissions to another acute care facility within one day of discharge, counting only the final admission in a transfer chain as the index hospitalization. A LAMA discharge followed by a readmission within one day was treated as index hospitalizations with a subsequent readmission, not as a transfer. Third, we did not follow AHRQ's recommendation of excluding patients with 20 or more hospitalizations because we believed it would exclude real patients with SCD.

The crude readmission rate was calculated as the percentage of index hospitalizations resulting in readmission. This approach ensures that multiple readmissions within a 30-day window are not double-counted, as each hospitalization can serve as only one index event.

---

[3] Agency for Healthcare Research and Quality (AHRQ). Methods: Calculating Readmissions for HCUPnet. Healthcare Cost and Utilization Project. Accessed May 2025.
https://hcup-us.ahrq.gov/tech_assist/data_query/HCUPnet_Readmissions_Methodology.pdf

**eTable 1:** Hospital-level leave against medical advice among adult sickle cell disease (SCD) hospitalizations, January 1, 2022-November 30, 2023
*Facilities (N=45) sorted by adjusted SCD LAMA using the baseline model*

| | | SCD LAMA – baseline model | | | | SCD LAMA - crude and adjusted rates excluding patients with 10 or more SCD hospitalizations | | | | SCD LAMA – baseline model plus patient-level random intercept | | |
|---|---|---|---|---|---|---|---|---|---|---|---|---|
| **Hospital** | **N SCD hosp.** | **Crude Rate** | **Adjusted Rate** | **CrI 95 (lower)** | **CrI 95 (upper)** | **Crude Rate** | **Adjusted Rate** | **CrI 95 (lower)** | **CrI 95 (upper)** | **Adjusted Rate** | **CrI 95 (lower)** | **CrI 95 (upper)** |
| **1** | 100-199 | 32.1 | 29.5 | 22.1 | 39.4 | 25.0 | 22.7 | 15.5 | 31.8 | 17.8 | 13.5 | 23.5 |
| **2** | 100-199 | 35.4 | 28.0 | 20.7 | 37.2 | 27.9 | 19.0 | 12.2 | 28.2 | 19.3 | 14.4 | 26.8 |
| **3** | 100-199 | 41.9 | 27.5 | 21.4 | 35.7 | 28.6 | 18.3 | 11.0 | 29.3 | 23.0 | 16.3 | 34.5 |
| **4** | 100-199 | 34.5 | 26.6 | 19.6 | 34.9 | 10.4 | 10.4 | 5.0 | 19.0 | 19.2 | 13.9 | 28.2 |
| **5** | 20-49 | 37.1 | 24.8 | 15.5 | 37.1 | 27.3 | 15.0 | 7.7 | 25.2 | 15.6 | 10.0 | 24.0 |
| **6** | 20-49 | 24.1 | 22.4 | 11.4 | 40.9 | 21.7 | 18.6 | 8.2 | 37.3 | 23.4 | 10.8 | 47.5 |
| **7** | 50-99 | 28.3 | 21.7 | 13.7 | 32.5 | 17.4 | 12.2 | 6.7 | 20.9 | 26.6 | 16.1 | 44.5 |
| **8** | 100-199 | 24.6 | 21.6 | 15.4 | 29.8 | 19.2 | 12.3 | 7.2 | 19.3 | 21.6 | 14.4 | 32.7 |
| **9** | 200-299 | 30.4 | 20.4 | 16.0 | 26.3 | 24.0 | 14.0 | 9.1 | 21.0 | 20.1 | 14.2 | 28.8 |
| **10** | 20-49 | 27.6 | 19.9 | 11.3 | 32.4 | 26.1 | 15.1 | 7.6 | 27.4 | 18.5 | 10.1 | 30.4 |
| **11** | 50-99 | 21.6 | 19.6 | 12.9 | 28.4 | 16.2 | 13.1 | 7.2 | 21.1 | 17.9 | 12.2 | 25.9 |
| **12** | 20-49 | 25.0 | 19.5 | 11.1 | 31.9 | 21.9 | 13.8 | 7.2 | 23.7 | 16.8 | 9.8 | 26.0 |
| **13** | 100-199 | 24.6 | 19.5 | 14.9 | 25.6 | 16.5 | 15.5 | 9.1 | 24.2 | 21.9 | 15.2 | 31.2 |
| **14** | 20-49 | 25.0 | 18.9 | 9.3 | 33.3 | 11.8 | 9.7 | 3.8 | 21.3 | 18.6 | 8.7 | 35.3 |
| **15** | 20-49 | 28.0 | 18.8 | 9.9 | 31.3 | 33.3 | 17.0 | 9.0 | 29.2 | 17.9 | 8.6 | 34.2 |
| **16** | 50-99 | 18.0 | 18.5 | 10.5 | 31.7 | 15.6 | 13.4 | 6.9 | 23.8 | 17.5 | 10.1 | 31.1 |
| **17** | 100-199 | 18.9 | 17.0 | 12.3 | 22.8 | 15.0 | 11.8 | 7.4 | 18.3 | 13.4 | 9.6 | 18.6 |
| **18** | 50-99 | 24.6 | 16.4 | 10.6 | 24.8 | 23.5 | 13.2 | 7.9 | 20.7 | 20.3 | 11.8 | 33.5 |
| **19** | 100-199 | 14.7 | 15.1 | 9.1 | 22.9 | 12.9 | 10.1 | 5.9 | 17.6 | 17.4 | 10.4 | 27.1 |
| **20** | 20-49 | 20.5 | 14.7 | 8.2 | 23.7 | 28.0 | 16.6 | 8.7 | 29.2 | 19.8 | 10.9 | 37.6 |
| **21** | 100-199 | 19.1 | 14.7 | 10.1 | 20.1 | 21.1 | 15.1 | 9.4 | 22.9 | 14.0 | 9.5 | 20.0 |
| **22** | 50-99 | 14.6 | 14.2 | 8.6 | 22.4 | 12.5 | 12.0 | 6.7 | 20.1 | 14.3 | 8.4 | 22.7 |
| **23** | 20-49 | 13.3 | 14.1 | 7.0 | 23.8 | 7.1 | 8.5 | 3.7 | 15.8 | 17.0 | 7.8 | 36.8 |
| **24** | 400-499 | 14.6 | 13.4 | 10.3 | 17.6 | 10.4 | 9.2 | 6.3 | 13.1 | 16.9 | 12.4 | 22.9 |

| | | | | | | | | | | | | |
|---|---|---|---|---|---|---|---|---|---|---|---|---|
| **25** | 100-199 | 14.8 | 13.3 | 8.8 | 19.0 | 13.6 | 11.4 | 6.6 | 17.8 | 12.5 | 8.4 | 18.5 |
| **26** | 20-49 | 9.5 | 13.3 | 5.2 | 28.2 | 10.0 | 10.6 | 4.3 | 21.8 | 12.9 | 3.5 | 38.7 |
| **27** | 100-199 | 13.0 | 13.1 | 8.3 | 19.4 | 11.7 | 10.8 | 6.4 | 16.9 | 13.8 | 8.7 | 22.1 |
| **28** | 50-99 | 17.3 | 12.7 | 7.8 | 19.1 | 16.9 | 10.5 | 6.2 | 16.0 | 13.5 | 8.7 | 20.7 |
| **29** | 50-99 | 8.7 | 11.7 | 5.9 | 20.0 | 2.3 | 6.0 | 2.4 | 13.2 | 9.5 | 5.0 | 17.6 |
| **30** | 100-199 | 9.4 | 11.2 | 6.7 | 17.9 | 8.9 | 8.6 | 4.3 | 15.3 | 12.0 | 6.7 | 21.1 |
| **31** | 300-399 | 13.4 | 11.2 | 7.9 | 15.1 | 16.1 | 11.8 | 8.2 | 17.0 | 15.2 | 10.7 | 21.8 |
| **32** | 400-499 | 9.3 | 10.9 | 7.9 | 14.7 | 6.5 | 7.6 | 4.9 | 11.5 | 10.8 | 7.6 | 15.8 |
| **33** | 200-299 | 11.7 | 10.8 | 7.3 | 15.7 | 12.8 | 10.1 | 6.1 | 15.8 | 10.7 | 6.7 | 16.6 |
| **34** | 200-299 | 13.6 | 10.2 | 7.0 | 14.2 | 10.2 | 8.4 | 4.9 | 14.1 | 11.6 | 7.3 | 17.6 |
| **35** | 100-199 | 7.6 | 9.8 | 5.4 | 15.7 | 4.7 | 6.4 | 3.0 | 11.8 | 8.5 | 5.0 | 13.6 |
| **36** | 100-199 | 9.0 | 9.6 | 6.0 | 14.7 | 4.6 | 5.7 | 2.7 | 10.3 | 7.9 | 4.7 | 12.4 |
| **37** | 100-199 | 10.1 | 9.1 | 5.7 | 14.9 | 25.5 | 17.5 | 10.3 | 26.2 | 14.8 | 8.1 | 26.2 |
| **38** | 20-49 | 0.0 | 8.7 | 2.6 | 22.7 | 0.0 | 7.9 | 2.2 | 23.2 | 9.5 | 1.3 | 56.5 |
| **39** | 200-299 | 9.0 | 8.7 | 6.0 | 12.7 | 8.6 | 7.6 | 4.3 | 12.7 | 7.9 | 5.1 | 12.0 |
| **40** | 700-799 | 9.6 | 8.4 | 6.4 | 10.9 | 6.4 | 6.3 | 4.2 | 9.1 | 7.7 | 5.7 | 11.0 |
| **41** | 600-699 | 7.8 | 7.8 | 5.6 | 10.6 | 4.2 | 4.7 | 3.0 | 7.2 | 8.2 | 6.1 | 11.1 |
| **42** | 300-399 | 6.3 | 7.5 | 4.8 | 11.1 | 6.2 | 6.1 | 3.8 | 9.9 | 7.9 | 4.9 | 12.3 |
| **43** | 400-499 | 6.1 | 6.6 | 4.5 | 9.4 | 4.0 | 4.7 | 2.9 | 7.3 | 7.2 | 5.0 | 10.2 |
| **44** | 300-399 | 5.6 | 6.1 | 3.9 | 8.8 | 4.5 | 5.3 | 3.1 | 8.5 | 7.7 | 4.9 | 12.7 |
| **45** | 50-99 | 1.1 | 4.5 | 1.8 | 8.5 | 0.0 | 4.1 | 1.6 | 9.0 | 4.2 | 1.5 | 10.1 |

Data source: New York State Department of Health, Statewide Planning and Research Cooperative System (SPARCS), Hospital Inpatient Discharges. See below for additional disclaimer.

**eTable 2:** Hospital-level leave against medical advice among adult all-cause hospitalizations, January 1, 2022-November 30, 2023
*Facilities (N=45) sorted by adjusted LAMA using the baseline model*

| | | All-cause LAMA - baseline model | | | | All-cause LAMA - crude and adjusted rates excluding patients with 10 or more hospitalizations | | | |
|---|---|---|---|---|---|---|---|---|---|
| **Hospital** | **N hosp.** | **Crude Rate** | **Adjusted Rate** | **CrI 95 (lower)** | **CrI 95 (upper)** | **Crude Rate** | **Adjusted Rate** | **CrI 95 (lower)** | **CrI 95 (upper)** |
| **27** | 10,000-19,999 | 9.4 | 8.9 | 7.7 | 10.4 | 8.7 | 8.2 | 6.9 | 9.5 |
| **26** | 10,000-19,999 | 9.7 | 8.0 | 6.9 | 9.3 | 8.6 | 7.0 | 6.0 | 8.2 |
| **31** | 10,000-19,999 | 10.0 | 7.6 | 6.6 | 8.9 | 9.3 | 7.0 | 6.0 | 8.2 |
| **17** | 30,000-39,999 | 10.4 | 7.5 | 6.5 | 8.7 | 9.4 | 6.9 | 5.9 | 8.0 |
| **7** | 10,000-19,999 | 8.3 | 6.6 | 5.7 | 7.8 | 7.8 | 6.1 | 5.2 | 7.2 |
| **11** | 30,000-39,999 | 4.8 | 6.5 | 5.7 | 7.7 | 4.2 | 5.6 | 4.8 | 6.6 |
| **21** | 20,000-29,999 | 9.3 | 6.4 | 5.6 | 7.5 | 8.5 | 5.9 | 5.1 | 6.9 |
| **4** | 10,000-19,999 | 6.3 | 6.3 | 5.4 | 7.5 | 5.2 | 5.1 | 4.3 | 6.1 |
| **34** | < 10,000 | 10.9 | 6.1 | 5.3 | 7.1 | 10.3 | 5.6 | 4.8 | 6.6 |
| **2** | 10,000-19,999 | 9.2 | 6.1 | 5.2 | 7.1 | 8.2 | 5.4 | 4.6 | 6.4 |
| **3** | 10,000-19,999 | 8.1 | 6.0 | 5.2 | 7.0 | 7.3 | 5.4 | 4.6 | 6.4 |
| **37** | 20,000-29,999 | 6.2 | 5.7 | 4.9 | 6.6 | 6.1 | 5.4 | 4.6 | 6.4 |
| **18** | 10,000-19,999 | 7.0 | 5.6 | 4.8 | 6.6 | 6.8 | 5.3 | 4.5 | 6.2 |
| **28** | 20,000-29,999 | 10.2 | 5.6 | 4.8 | 6.5 | 9.5 | 5.1 | 4.4 | 5.9 |
| **39** | 10,000-19,999 | 5.0 | 5.5 | 4.6 | 6.5 | 4.3 | 4.7 | 3.9 | 5.5 |
| **1** | 20,000-29,999 | 4.9 | 5.3 | 4.6 | 6.2 | 4.3 | 4.7 | 3.9 | 5.5 |
| **16** | 20,000-29,999 | 6.0 | 4.8 | 4.2 | 5.6 | 5.4 | 4.3 | 3.6 | 5.0 |
| **24** | 20,000-29,999 | 5.8 | 4.8 | 4.1 | 5.6 | 5.1 | 4.1 | 3.5 | 4.9 |
| **13** | 20,000-29,999 | 6.0 | 4.7 | 4.1 | 5.5 | 5.4 | 4.3 | 3.6 | 5.0 |
| **8** | 10,000-19,999 | 3.8 | 4.6 | 3.8 | 5.5 | 3.5 | 4.0 | 3.3 | 4.7 |
| **9** | 40,000-49,000 | 5.1 | 4.1 | 3.6 | 4.8 | 4.1 | 3.4 | 2.9 | 4.0 |
| **6** | 10,000-19,999 | 5.2 | 3.9 | 3.3 | 4.6 | 4.7 | 3.6 | 3.0 | 4.2 |
| **14** | 10,000-19,999 | 5.7 | 3.8 | 3.2 | 4.5 | 4.9 | 3.3 | 2.8 | 3.9 |
| **25** | 20,000-29,999 | 3.8 | 3.7 | 3.2 | 4.3 | 3.4 | 3.4 | 2.9 | 4.0 |

| | | | | | | | | | |
|---|---|---|---|---|---|---|---|---|---|
| **20** | 20,000-29,999 | 3.1 | 3.7 | 3.2 | 4.4 | 2.7 | 3.2 | 2.7 | 3.7 |
| **44** | 70,000-79,999 | 2.1 | 3.7 | 3.2 | 4.3 | 1.8 | 3.1 | 2.6 | 3.6 |
| **19** | 20,000-29,999 | 4.4 | 3.6 | 3.1 | 4.3 | 4.0 | 3.3 | 2.8 | 3.9 |
| **36** | 40,000-49,000 | 3.0 | 3.5 | 3.0 | 4.2 | 2.7 | 3.1 | 2.7 | 3.7 |
| **10** | 20,000-29,999 | 4.7 | 3.5 | 3.0 | 4.1 | 4.3 | 3.1 | 2.7 | 3.7 |
| **15** | 20,000-29,999 | 4.2 | 3.5 | 3.0 | 4.1 | 3.7 | 3.1 | 2.7 | 3.7 |
| **5** | 40,000-49,000 | 2.3 | 3.4 | 2.9 | 4.0 | 2.1 | 3.0 | 2.5 | 3.5 |
| **32** | 40,000-49,000 | 2.6 | 3.2 | 2.8 | 3.8 | 2.3 | 2.8 | 2.3 | 3.2 |
| **40** | 70,000-79,999 | 2.3 | 3.1 | 2.6 | 3.6 | 2.1 | 2.7 | 2.3 | 3.2 |
| **22** | 40,000-49,000 | 2.5 | 3.1 | 2.6 | 3.6 | 2.4 | 2.7 | 2.3 | 3.2 |
| **41** | 50,000-59,000 | 2.9 | 2.9 | 2.5 | 3.4 | 2.6 | 2.6 | 2.2 | 3.0 |
| **29** | 10,000-19,999 | 2.9 | 2.9 | 2.5 | 3.5 | 2.6 | 2.6 | 2.1 | 3.1 |
| **12** | 10,000-19,999 | 2.7 | 2.7 | 2.3 | 3.1 | 2.6 | 2.5 | 2.1 | 3.0 |
| **23** | 40,000-49,000 | 2.3 | 2.6 | 2.3 | 3.1 | 2.2 | 2.4 | 2.0 | 2.8 |
| **33** | 30,000-39,999 | 2.4 | 2.6 | 2.2 | 3.1 | 2.0 | 2.2 | 1.8 | 2.6 |
| **30** | 40,000-49,000 | 2.1 | 2.5 | 2.1 | 3.0 | 2.0 | 2.2 | 1.9 | 2.6 |
| **43** | 60,000-69,999 | 2.0 | 2.5 | 2.1 | 3.0 | 1.8 | 2.2 | 1.8 | 2.6 |
| **45** | 10,000-19,999 | 2.0 | 2.2 | 1.9 | 2.7 | 1.9 | 2.0 | 1.7 | 2.4 |
| **35** | 60,000-69,999 | 1.4 | 2.2 | 1.9 | 2.6 | 1.2 | 1.8 | 1.5 | 2.1 |
| **42** | 50,000-59,000 | 1.4 | 1.7 | 1.5 | 2.0 | 1.2 | 1.4 | 1.2 | 1.7 |
| **38** | 30,000-39,999 | 0.3 | 0.6 | 0.4 | 0.7 | 0.3 | 0.5 | 0.4 | 0.6 |

Data source: New York State Department of Health, Statewide Planning and Research Cooperative System (SPARCS), Hospital Inpatient Discharges. See below for additional disclaimer.

**eTable 3:** Hierarchical logistic regression models predicting 30-day readmissions in sickle cell disease (SCD) hospitalizations: comparing leave against medical advice (LAMA) discharges vs. non-LAMA discharges

| | **Model (0)** N= 7,939 | **Model (1)** N = 7,925 | **Model (2)** N=7,896 |
|---|---|---|---|
| | *Unadjusted odds ratio [CI 95]* | *Adjusted odds ratio [CI 95]* | *Adjusted odds ratio [CI 95]* |
| **Discharge status** | | | |
| Non-LAMA [Reference] | 1 | 1 | 1 |
| LAMA | **1.94 [1.58, 2.38]*** ** | **1.63 [1.39, 1.91]*** ** | **1.61 [1.37, 1.89]*** ** |
| **Patient characteristics** | | | |
| Female [Reference] | | 1 | 1 |
| Male | | 1.33 [1.08, 1.64]** | 1.46 [1.22, 1.74]*** |
| Age 18-29 [Reference] | | 1 | 1 |
| Age 30-49 | | 1.15 [0.92, 1.42] | 1.09 [0.89, 1.32] |
| Age 50-65 | | 0.78 [0.53, 1.15] | 0.73 [0.51, 1.04] |
| Age 65+ | | 0.53 [0.22, 1.29] | 0.45 [0.20, 0.98]* |
| Non-ED admission [Reference] | | 1 | 1 |
| ED admission | | 1.42 [0.99, 2.05] | 1.2 [0.85, 1.68] |
| Severity of illness present on admission 1 (lowest) [Reference] | | 1 | 1 |
| Severity of illness present on admission 2 | | 0.85 [0.74, 0.97]* | 0.84 [0.74, 0.96]* |
| Severity of illness present on admission 3 | | 0.87 [0.75, 1.01] | 0.83 [0.72, 0.96]* |
| Severity of illness present on admission 4 (highest) | | 0.58 [0.41, 0.82]** | 0.58 [0.41, 0.82]** |
| Elixhauser comorbidity fixed effects | | Yes | Yes |
| **Other patient demographics** | | | |
| Non-Hispanic White [Reference] | | | 1 |
| Asian or Pacific Islander | | | 0.96 [0.22, 4.13] |
| Black | | | 1.7 [0.44, 6.57] |
| Hispanic | | | 1.34 [0.32, 5.61] |

| | | | |
|---|---|---|---|
| Other race or ethnicity | | | 0.97 [0.25, 3.83] |
| Private insurance [Reference] | | | 1 |
| Medicaid | | | 2.09 [1.64, 2.66]*** |
| Medicare | | | 2.35 [1.62, 3.41]*** |
| Other insurance | | | 1.64 [0.80, 3.35] |
| Self-pay | | | 0.71 [0.44, 1.14] |
| Neighborhood fixed effects | | | Yes |
| **Hospital random intercept** | | | |
| Hospital random intercept (variance) | | .21 [.13-.34] | .19 [.10-.34] |
| Intra-cluster correlation coefficient (ICC) | | 6.1% | 5.4% |
| Groups (hospitals) | | 45 | 45 |

* p< .05, ** p< .01, *** p<.001.
Model (1) adjusted for patient characteristics and model (2) added demographic and neighborhood predictors.
Less than 1% of observations were not used due to missing data for severity of illness, sex, or neighborhood variables.
Data source: New York State Department of Health, Statewide Planning and Research Cooperative System (SPARCS), Hospital Inpatient Discharges. See below for additional disclaimer.

**eTable 4:** Hierarchical logistic regression models predicting 30-day readmissions in all-cause hospitalizations: comparing leave against medical advice (LAMA) discharges vs. non-LAMA discharges

| | **Model (0)** N= 1,358,673 | **Model (1)** N = 1,357,736 | **Model (2)** N=1,357,716 |
|---|---|---|---|
| | *Unadjusted odds ratio [CI 95]* | *Adjusted odds ratio [CI 95]* | *Adjusted odds ratio [CI 95]* |
| **Discharge status** | | | |
| Non-LAMA [Reference] | 1 | 1 | 1 |
| LAMA | **2.51 [2.28, 2.76]***** | **2.10 [1.96, 2.26]***** | **2.06 [1.93, 2.21]***** |
| **Patient characteristics** | | | |
| Female [Reference] | | 1 | 1 |
| Male | | 1.12 [1.09, 1.14]*** | 1.14 [1.11, 1.16]*** |
| Age 18-29 [Reference] | | 1 | 1 |
| Age 30-49 | | 1.03 [0.97, 1.09] | 1.05 [1.00, 1.11] |
| Age 50-65 | | 1.06 [0.98, 1.14] | 1.08 [1.01, 1.16]* |
| Age 65+ | | 0.94 [0.87, 1.02] | 0.93 [0.85, 1.01] |
| Non-ED admission [Reference] | | 1 | 1 |
| ED admission | | 1.52 [1.39, 1.66]*** | 1.45 [1.35, 1.57]*** |
| Severity of illness present on admission = 1 (lowest) [Reference] | | 1 | 1 |
| Severity of illness present on admission = 2 | | 1.5 [1.44, 1.55]*** | 1.48 [1.43, 1.53]*** |
| Severity of illness present on admission = 3 | | 1.95 [1.84, 2.06]*** | 1.92 [1.81, 2.04]*** |
| Severity of illness present on admission = 4 (highest) | | 2.12 [1.96, 2.29]*** | 2.11 [1.94, 2.28]*** |
| Elixhauser comorbidity fixed effects | | Yes | Yes |
| **Other patient demographics** | | | |
| Non-Hispanic White [Reference] | | | 1 |
| Asian or Pacific Islander | | | 0.97 [0.92, 1.02] |
| Black | | | 1.13 [1.08, 1.17]*** |
| Hispanic | | | 1.04 [0.99, 1.10] |
| Other race or ethnicity | | | 0.92 [0.88, 0.97]*** |

| | | | |
|---|---|---|---|
| Private insurance [Reference] | | | 1 |
| Medicaid | | | 1.42 [1.34, 1.50]*** |
| Medicare | | | 1.35 [1.27, 1.44]*** |
| Other insurance | | | 1.03 [0.88, 1.20] |
| Self-pay | | | 0.9 [0.82, 0.98]* |
| Neighborhood fixed effects | | | Yes |
| **Hospital random intercept** | | | |
| Hospital random intercept (variance) | | .034 [.022-.053] | .030 [.020-.045] |
| Intra-cluster correlation coefficient (ICC) | | 1% | < 1% |
| Groups (hospitals) | | 45 | 45 |

* p< .05, ** p< .01, *** p<.001.
Model (1) adjusted for patient characteristics and model (2) added demographic and neighborhood predictors.
Less than 1% of observations were not used due to missing data for severity of illness, sex, or neighborhood variables.
Data source: New York State Department of Health, Statewide Planning and Research Cooperative System (SPARCS), Hospital Inpatient Discharges.

**Data disclaimer for SPARCS**

The raw SPARCS data used to produce this publication was provided by the New York State Department of Health. However, the calculations, metrics, conclusions derived, and views expressed herein are those of the author(s) and do not reflect the work, conclusions, or views of New York State. The New York State Department of Health, its employees, officers, and agents make no representation, warranty or guarantee as to the accuracy, completeness, currency, or suitability of the information provided here.
